# Estimation of Abortions in the Province of Buenos Aires, Argentina: a Bayesian Approach

**DOI:** 10.1101/2023.07.31.23293446

**Authors:** Andrea Paz, Sharon Josid, María Carla Rodríguez, Giselle Lamela, Matías Poullain, Lupe Marín, Franco Marsico

**Affiliations:** Ministerio de Salud, Provincia de Buenos Aires, Buenos Aires, Argentina; Facultad de Ciencias Exactas y Naturales, Universidad de Buenos Aires, Buenos Aires, Argentina

## Abstract

Abortion is one of the reasons for hospital discharge among women of fertile age in Argentina. The criminalization of abortion prior to the enactment of the Voluntary Interruption of Pregnancy Act (IVE), coupled with fragmentation of the health system, has hindered the availability of reliable records to quantify induced abortions in the country. Given the lack of information reflecting the magnitude of these practices, it is necessary to perform an accurate estimation to characterize abortion access within the healthcare system. Using a Bayesian approach and abortion records in the province of Buenos Aires in 2021, the incidence of abortions per thousand women of fertile age was estimated. This study, based on a direct case count methodology, is the first of its kind in Argentina. The method used allows for the correction of bias caused by under-reporting of practices. An incidence of 29.92 (Credibility Interval, CI95: [29.38; 30.47]) abortions per thousand women of fertile age was obtained during 2021. The results are consistent with previous national research and agree with estimates made for the region. In conclusion, the estimates of abortion incidence highlight its frequency in the lives of people with gestational capacity, underlining the importance of fully guaranteeing the right to comprehensive sexual health through a wide network of abortion access.

## 1 Introduction

Abortion is a frequent event in the lives of individuals with reproductive capacities; particularly in Argentina, it is one of the important reasons for hospital discharge in women of fertile age [1]. The legal framework surrounding abortion practices was recently modified in Argentina. After 17 years of the Campaign for Legal, Safe and Free Abortion, in December 2020, the National Law N° 27.610 of Voluntary Interruption of Pregnancy was sanctioned (IVE Law) [2]. Before this Law, the National Penal Code criminalised abortion, with some exceptions indicated in its article Nro 86 [3]: (i) risk to the life and health of the pregnant person or (ii) if the pregnancy was the result of a rape. Although pregnancy interruption for these causes has been in effect since 1921, access was met with multiple obstacles in the health system [4, 5].

The IVE Law of 2020 opens a new normative framework, as it replaces the system of causes established in the Penal Code since 1921 by recognizing the right of pregnant people to voluntarily interrupt their pregnancy up to the fourteenth week inclusive of the gestational process [2]. Outside of this term, the pregnant person has the right to access the legal interruption of pregnancy if it was the result of a rape or if the life or health of the pregnant person was in danger/risk. The effective implementation of abortion access that the IVE Law carries special difficulties throughout Argentina, due to territorial heterogeneities and inequalities, levels of government involved, fragmentation of the health system [6], and the historical tensions and disputes of meaning that abortion practice produces.

The Argentine health system is a fragmented system in three subsectors: public, social security, and private. In turn it has different levels of government in the case of the public subsector, finding health establishments of different dependencies: municipal, provincial, or national [6]. This fragmentation in the province of Buenos Aires has special complexity since it is made up of 135 municipalities or local governments, on whom almost all primary care centers and 218 municipal hospitals depend [7]. This decentralization of primary health care to the local level [8] adds greater complexity to the system, increased by inequalities between municipalities and within them. The province has 95 hospitals under its dependence, along with 5 national hospitals, of the second and third level of care. Also, to organize the health management of the territory, it is divided into twelve sanitary regions that are distributed in management and government areas integrated at the central level, from which health policies are carried out from an intermediate position between the provincial and municipal level. Buenos Aires is the most populated province in Argentina. In 2021, the estimated population was 17,709,598 inhabitants [9], representing 38.6% of the total population of the country. The distribution is uneven, concentrating the largest amount of population in the 40 municipalities of the Buenos Aires Metropolitan Area (AMBA).

The criminalization of the abortion practice before the sanction of the IVE Law, together with the fragmentation of the health system, have made it difficult to have reliable records to quantify the events of induced abortions in the province of Buenos Aires. Atucha and Pailes [10] and Mario and Pantelides [11] made estimates for Argentina using indirect methodologies. These publications were made before the sanction of the IVE Law, so one of the main difficulties was the non-existence of records or unreliable records.

In the absence of information that indicates the magnitude of abortion in the province of Buenos Aires, it is necessary to accurately estimate it to characterise access to abortion in the health system. In 2020, Buenos Aires Ministry of Health developed the first centralised registry of abortions, making it possible to have data to make direct estimates of them, as has been done in similar studies in other countries [12, 13]. Moreover, the information healthcare system proved to be robust enough for monitoring real-world vaccination campaigns [14, 15, 16, 17, 18, 19, 20]. This study made an estimate of the number of abortions produced during the year 2021 in the public health subsector and throughout the province, based on direct methods. From the bibliographic search carried out, this would be the first estimate developed in the country based on the direct record of practices.

## 2 Methods

### 2.1 Systematic literature search

An exhaustive search of antecedents was carried out in the virtual libraries of scientific publications Scielo, PubMed, and Virtual Health Library (BVS) with the following steps: (i) The search was located in title and abstract using the reference terms “estimate” and “abortion”. Only in the case of the BVS to specify the initial search, a new filter was applied in the main subject (induced abortion/legal abortion), since more than 7,000 publications were obtained. (ii) A reading of the abstracts was carried out. Those publications that specifically referred to induced or voluntary abortions in humans were selected. Primarily it was evaluated that they were related to the calculation of incidence. (iii) Finally, the country or region of the publications was regionalized.

### 2.2 Data

As previously stated, the province is politically divided into 135 municipalities, grouped into 12 sanitary regions (Figure 1) as part of the political-administrative structure of the provincial Ministry of Health. From the last available national census [21], population information (total number and by five-year age groups) estimated for 2021 and information on the percentage of households with Unsatisfied Basic Needs (UBN) were obtained as a socioeconomic indicator for each municipalities.

**Figure 1:**
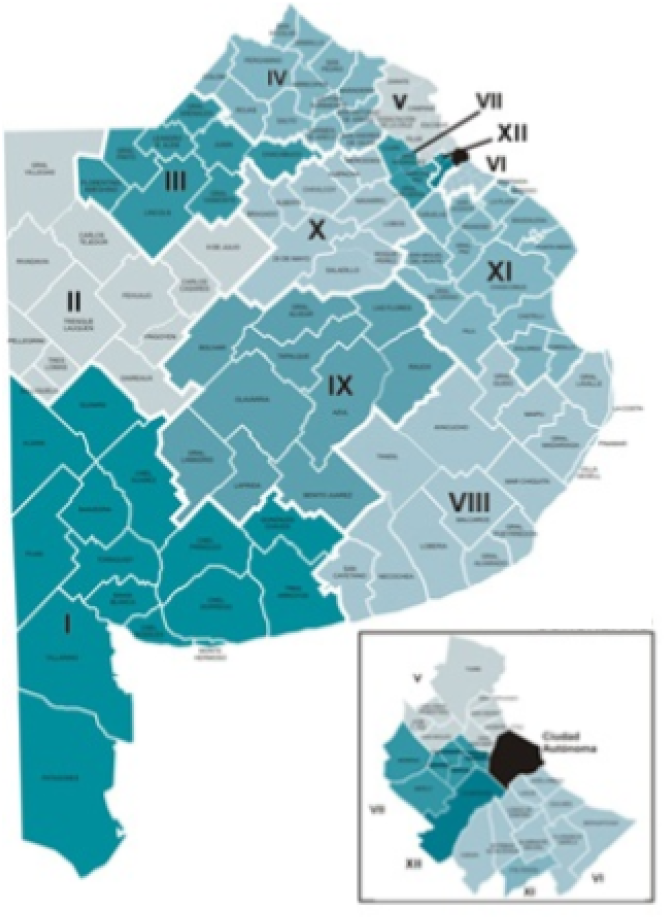
Sanitary regions of the Province of Buenos Aires.

The target population of this study were women of fertile age residing in the province of Buenos Aires who attend the public health subsector. By “woman of fertile age” is understood any person who identifies in the health records as female between the ages of 15 and 49 inclusive. It is estimated that in 2021 there are 4,361,010 women of fertile age residing in the province, of which 33.3% are cared for in the public subsector (1,450,700 women) [22]. However, there are significant heterogeneities in the territory, since while in some municipalities the proportion of women who have exclusive public coverage reaches 43.5%, in others it does not exceed 20.0%.

Table 1 shows the estimated number of women of fertile age by sanitary region and the unequal distribution of the number of health establishments that guarantee access to abortion.

**Table 1:** Descriptive statistics by sanitary region of health establishments and women of fertile age. Year 2021. Health facilities that guarantee abortion access for each region, number of womens in fertile age and the Rate of health facilities per 1000 womens.

| Region | Health facilities | Women (15-49 years old) | Rate |
| --- | --- | --- | --- |
| I | 36 | 161.268 | 0.22 |
| II | 17 | 62.015 | 0.27 |
| III | 8 | 58.437 | 0.14 |
| IV | 17 | 144.595 | 0.12 |
| V | 83 | 882.989 | 0.09 |
| VI | 95 | 1.057.049 | 0.09 |
| VII | 63 | 622.748 | 0.10 |
| VIII | 66 | 291.994 | 0.23 |
| IX | 34 | 72.853 | 0.47 |
| X | 15 | 77.289 | 0.19 |
| XI | 44 | 333.420 | 0.13 |
| XII | 15 | 596.353 | 0.03 |

Data on the number of abortions were obtained from the Unified Registry of IVE/ILE practices and use of misoprostol developed by the Ministry of Health of the province of Buenos Aires. It has information on 34,354 abortions performed in the province within the public health subsector during the year 2021. The registry is anonymous, recording the place where the practice was performed, the methodology used (surgery, pharmaceutical, etc.), the municipality of residence, and age. Also, analyzed data were aggregated at municipality level. Moreover, public information from the Non-Governmental Organisation “Socorristas en Red” was available. It is responsible for accompanying abortion situations. During 2021 they accompanied a total of 2,648 cases, of which 1,778 were abortions carried out within the community health system (self-managed). This implies that part of the population, having the public health subsector, performs practises outside it, in a self-managed way with the accompaniment of other networks. For estimation, this implies a source of under-registration, of the same nature as was observed in similar studies [12, 13, 27, 28]. The 1,778 cases represent 5.84% of the registered abortions. It is therefore taken that this is the lower bound of the under-registration (since there are also other NGOs, and self-managed abortions that are not registered). All proportions of patients by age within each sanitary region or type of health institution were compared. These comparisons were made pairwise with proportional Z tests [25] and with adjustment of p-values with the Holm method [26]. The values “No data” were not taken into account.

### 2.3 Statistical model

To generate estimates of the incidence of abortions in the population of women of fertile age taking into account the sources of uncertainty already mentioned, the following statistical approach was used:

First, the incidence of abortions per 1,000 women of fertile age was calculated for each sanitary region and for the entire province from formulas (1) and (2):

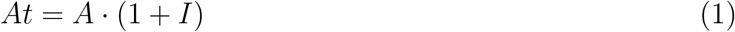

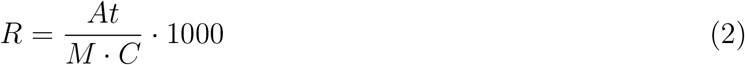

Where *At* is the total number of abortions, *A* is the registered number of abortions performed in the public subsector, *I* is the estimated proportion of abortions performed outside the formal health field, *M* is the projected number of women of fertile age residents for the year 2021, and *C* is the proportion with exclusive public health coverage. On the other hand, *R* is the incidence of abortions per 1,000 women of fertile age. This calculation was made for each municipality, for each sanitary region, and for the entire province.

Secondly, a *Direct* method was applied, where an interval for the incidence of abortions was estimated per 1,000 women of fertile age was estimated based on the estimated interval of the proportion of abortions performed outside the formal health field (6-10%) and formula (1). The range of the proportion of abortions performed outside the formal field is based on the data provided by local NGOs and international literature [12, 13, 27, 28].

Then, two similar Bayesian binomial regression models were performed. In both, the number of abortions was modeled based on the percentage of households with UBN [29] and the number of establishments in the public health field per 1,000 women of fertile age, taking the municipality as the unit of analysis. The formula for the linear predictor of the models is defined below (3).

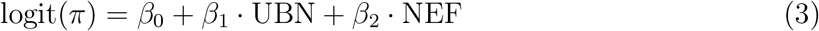

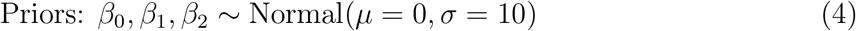

Where *π* is the probability that a woman has an abortion, and *β*_0_, *β*_1_ and *β*_2_ are the regression parameters corresponding to the intercept, the percentage of households with UBN, and the NEF (Number of Establishments per 1,000 women of fertile age), respectively.

For all these parameters, the prior distributions chosen were weakly informative since it was sought that the posterior distributions obtained are primarily defined by the data, since the previous information in relation to these parameters is scarce. With that same base model, two different models were trained, one with all the municipalities of the province and another only with the municipalities that belong to the sanitary regions with reliable registering, based on experts definition. These regions were VI, VIII, X, and XI, indicated as those where the Unified Registry of IVE/ILE was extensively applied. These models were called the *All Departments model* and the *Reliable Registering model*. With them, two interval estimates of R were made for each each sanitary region, and for the entire province. Then the three estimates (direct model, all departments model, and reliable registration rate model) were compared for the sanitary regions and the province. This sensitivity analysis allows for a clearer picture of the influence of uncertainty sources on the final estimate of the incidence of abortions. The R software, version 4.2.1 was used for data manipulation, statistical analysis, and visualisation. Specifically, for Bayesian modelling, the “bmrs” package [30] was used.

## 3 Results

In the literature search, a total of 1,343 scientific publications were found related to abortion incidence estimates, with 836 found in PubMed, 56 in Scielo, and 451 in the BVS. Of all the publications, 128 were specifically related to the calculation of incidence (Supplementary Table 1). There were 32 publications referring to abortion estimates in Latin American countries: Brazil (n= 13), Chile (n= 5), Colombia (n= 5), Mexico (n= 4), Cuba (n= 2), Peru (n= 2), Costa Rica (n= 1), Guatemala (n= 1) and Dominican Republic (n= 1). Of the total articles, only two present abortion estimates in Argentina, while none present disaggregated data for the province of Buenos Aires. Figure 1 shows the number of publications found per year.

Based on the event registry and during the period analysed, 34,354 abortions were recorded, of which 34,073 were performed in the public health sector with a residence in a municipality of the Province of Buenos Aires (PBA), while 281 were indicated within the PBA, but without specifying a municipality. One in every two abortions were performed in primary health care centres (CAPS), which implement promotion and prevention programmes, while more than eight out of every ten used pharmacological methods (Figure 2). Regarding the analysis by age groups, it was observed that 54.1% of the procedures were performed on women between 20 and 29 years old (Table 2). The age group distribution for the different sanitary regions did not show statistically significant differences (p-value > 0.05), except in the case of sanitary region VII, where the proportion of people aged 15 to 19 (13.4%) shows significant differences compared to the rest of the age groups (15.2%-19.0%, p-values<0.001) excluding the 45-49 age segment. Furthermore, in sanitary region VII, significant differences are also observed between the 20-24 age group (15.2%) in relation to the 25-29 year interval (16.9%, p-value = 0.02), 35-39 years (17.4%, p-value = 0.04) and 40-44 years (19.0%, p-value = 0.011).

**Table 2:**
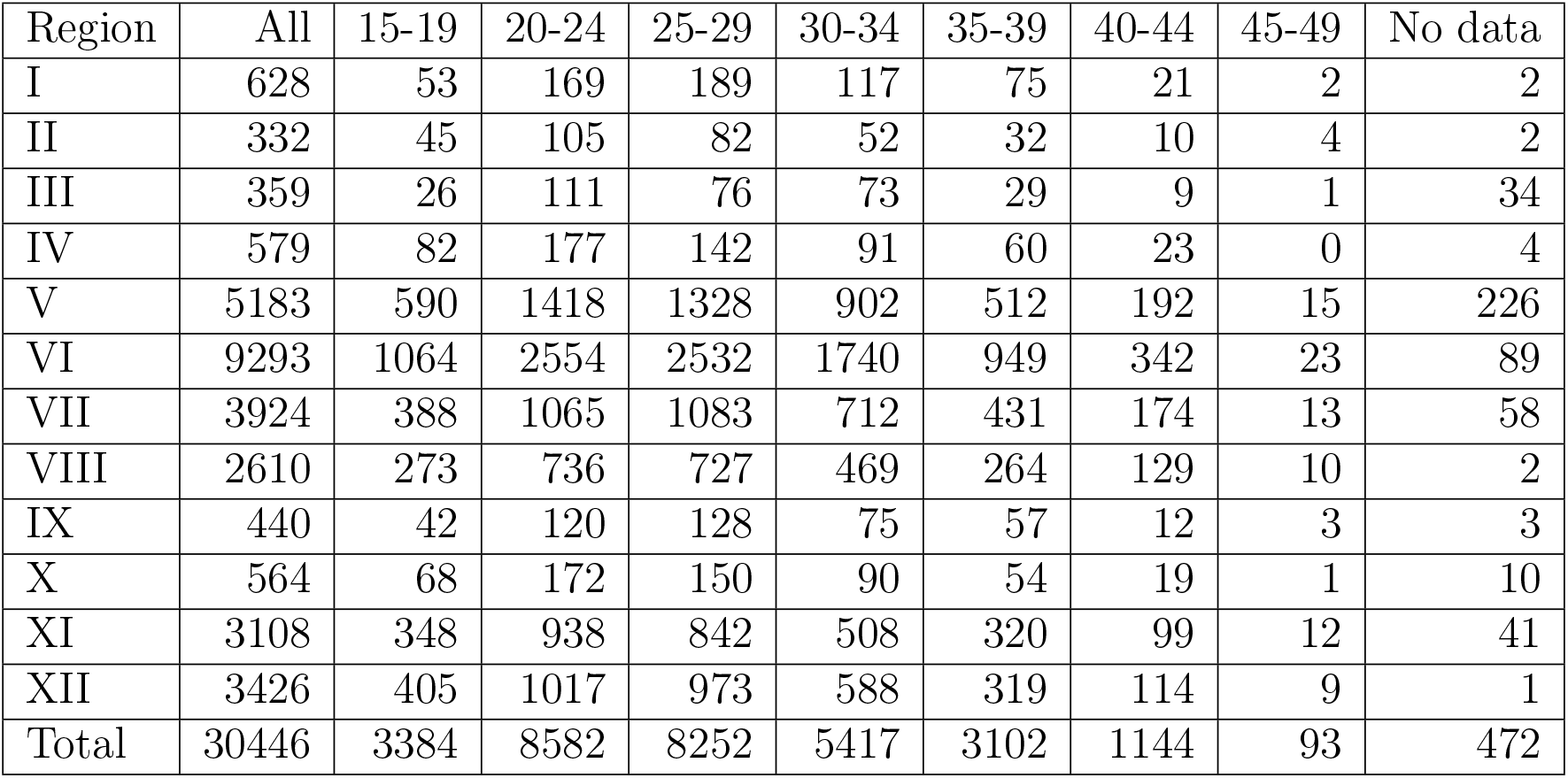
Number of registered abortions per age and sanitary region.

| Region | All | 15-19 | 20-24 | 25-29 | 30-34 | 35-39 | 40-44 | 45-49 | No data |
| --- | --- | --- | --- | --- | --- | --- | --- | --- | --- |
| I | 628 | 53 | 169 | 189 | 117 | 75 | 21 | 2 | 2 |
| II | 332 | 45 | 105 | 82 | 52 | 32 | 10 | 4 | 2 |
| III | 359 | 26 | 111 | 76 | 73 | 29 | 9 | 1 | 34 |
| IV | 579 | 82 | 177 | 142 | 91 | 60 | 23 | 0 | 4 |
| V | 5183 | 590 | 1418 | 1328 | 902 | 512 | 192 | 15 | 226 |
| VI | 9293 | 1064 | 2554 | 2532 | 1740 | 949 | 342 | 23 | 89 |
| VII | 3924 | 388 | 1065 | 1083 | 712 | 431 | 174 | 13 | 58 |
| VIII | 2610 | 273 | 736 | 727 | 469 | 264 | 129 | 10 | 2 |
| IX | 440 | 42 | 120 | 128 | 75 | 57 | 12 | 3 | 3 |
| X | 564 | 68 | 172 | 150 | 90 | 54 | 19 | 1 | 10 |
| XI | 3108 | 348 | 938 | 842 | 508 | 320 | 99 | 12 | 41 |
| XII | 3426 | 405 | 1017 | 973 | 588 | 319 | 114 | 9 | 1 |
| Total | 30446 | 3384 | 8582 | 8252 | 5417 | 3102 | 1144 | 93 | 472 |

**Figure 2:**
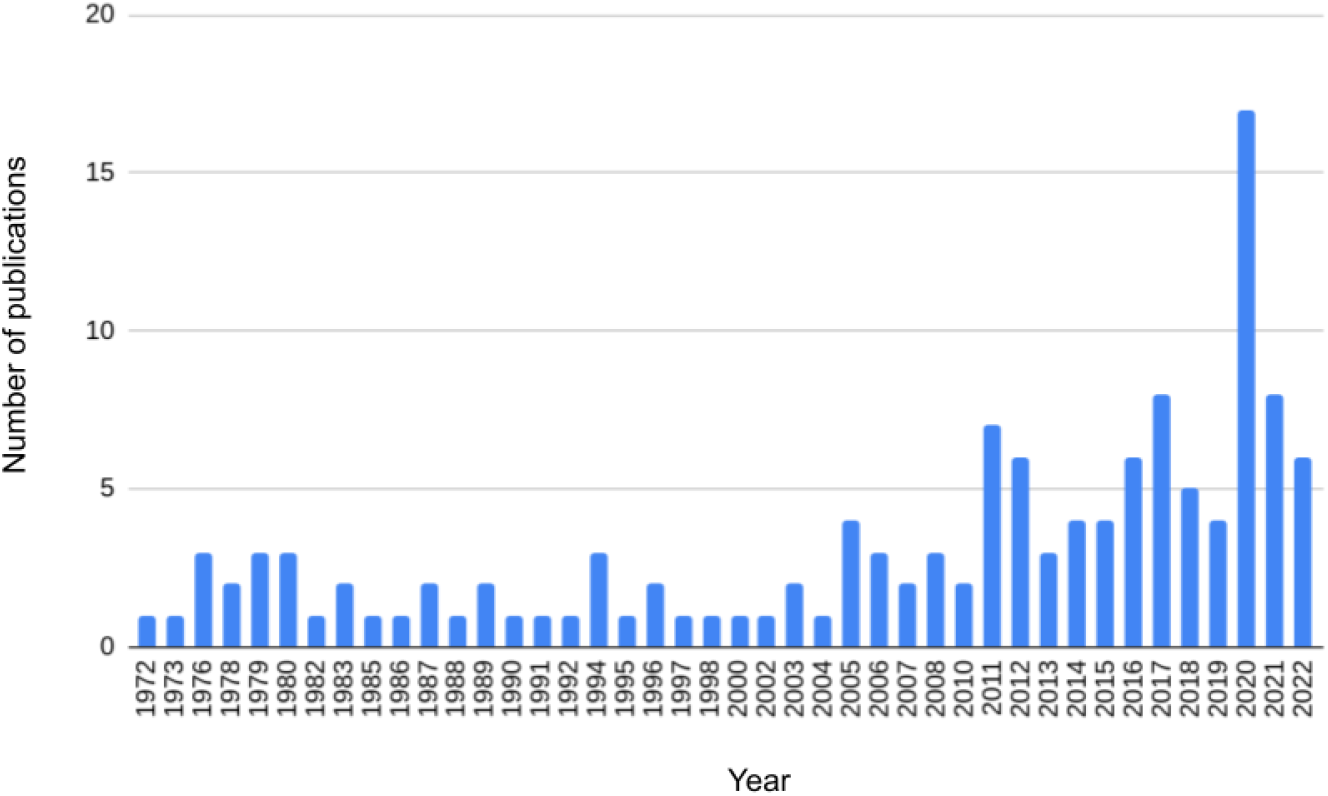
Number of scientific publications per year. 1972-2022.

In terms of the establishment where the procedures were performed, there are significant differences between the proportions of people aged 15 to 19 who attended the CAPS (46.9%) and provincial hospitals (30.9%) compared to the 20-24 group (50.8%, p-value = 0.01 and 27.3%, p-value < 0.01), 25-29 (52.0%, p-value < 0.01 and 26.3%, p-value < 0.01) and 30-34 (52.3%, p-value < 0.01 and 26.2%, p-value < 0.01). On the other hand, the 45-49 age range has a lower proportion of abortions performed in CAPS (42.7%) than the younger age groups (46.9%-52.3%) and a higher proportion of procedures performed in municipal hospitals (21.8%) compared to the rest of the younger groups (16.3%-17.1%). However, none of these differences is statistically significant.

The estimates of abortions per 1,000 women of fertile age and their 95% Credibility Intervals (IC95) by sanitary region are represented in Figure 3. In this figure, it can be observed that the Direct method retains the most variation of the estimates between the sanitary regions and that the estimates of the other two methods are usually higher. In the whole province, it was estimated by the Direct model that the number of abortions performed per 1,000 women of fertile age is between 25.50 and 28.05, while by the All Departments model, the estimated mean value was 26.45 (IC95: [26.08;26.83]) and by the Reliable Registering Rate model it was 29.92 (IC95: [29.38;30.47]). Based on these results, the total number of abortions performed in the province could be estimated. Assuming that the abortion rate between the private and public sectors is homogeneous, between 108,259 abortions (considering the lower value of the interval in the Direct model) and 129,374 (considering the higher value of the IC95 Reliable Registering model) are estimated for the province.

**Figure 3:**
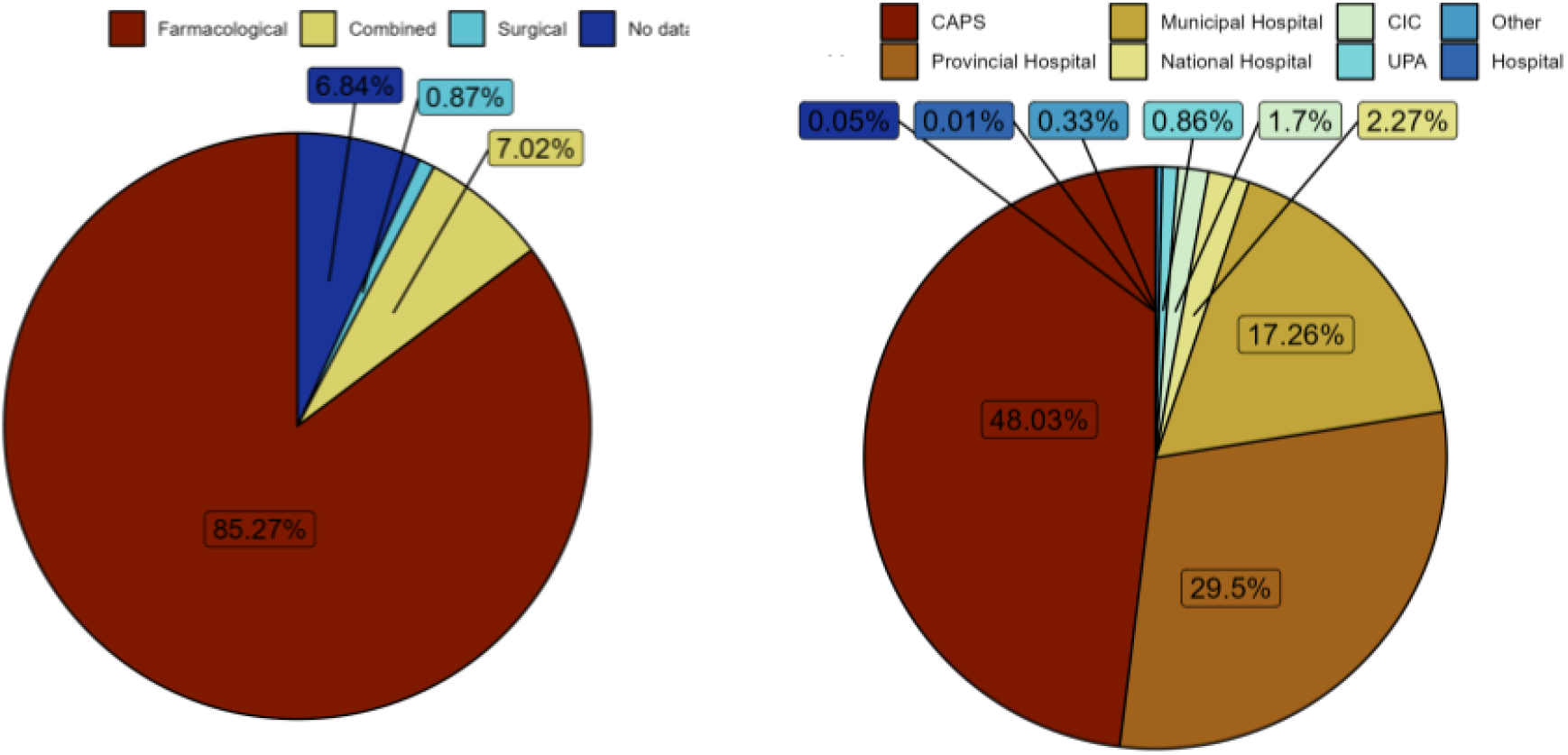
Pie charts of the proportion of registered abortions according to A) the type of institution where they were performed and B) the medical procedure.

**Figure 4:**
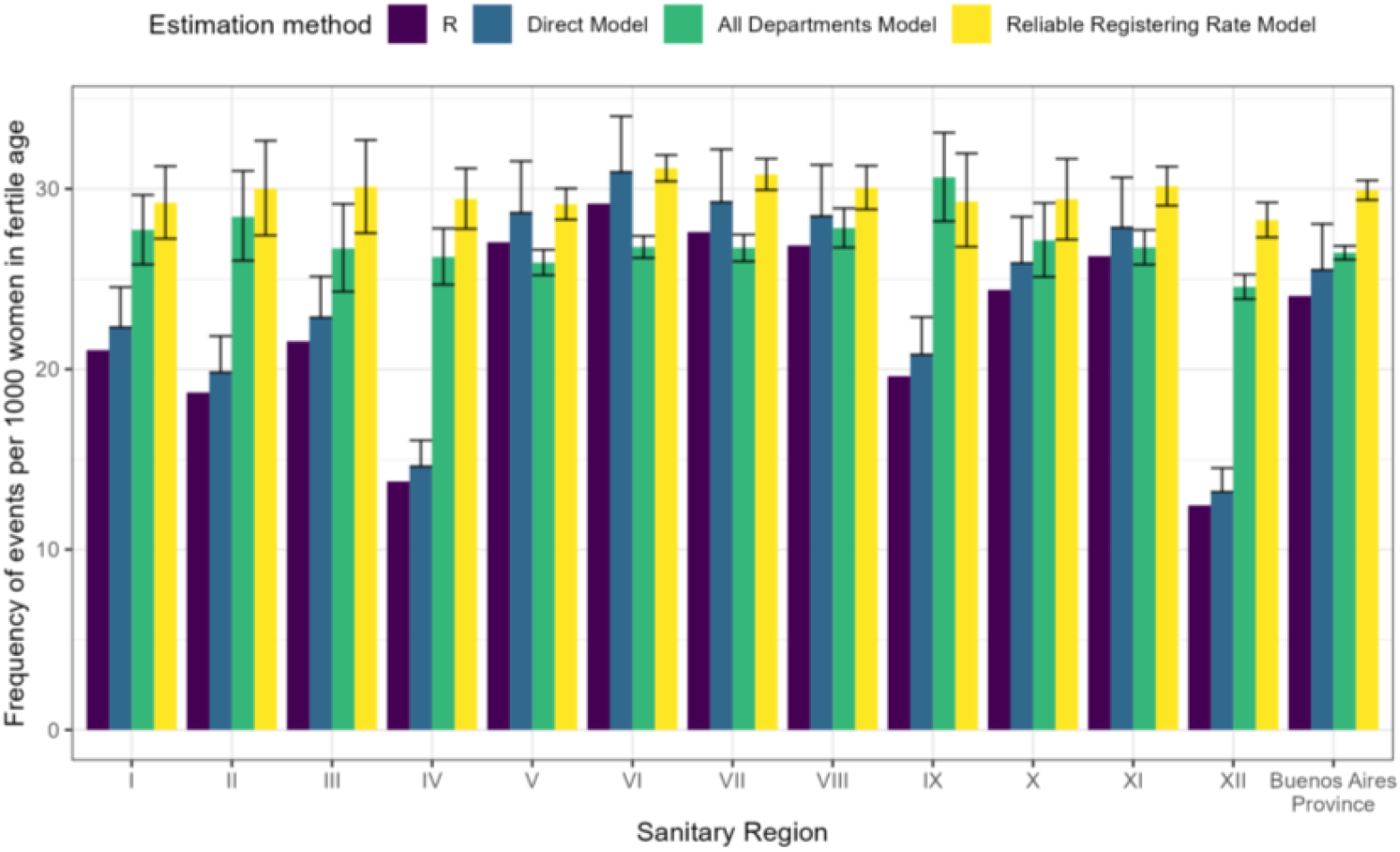
Bar chart of the estimated R value of abortions per 1,000 women of fertile age, their different estimates and their credibility intervals by sanitary region and for the province of Buenos Aires.

## 4 Discussion

To the best of our knowledge, this is the first study in Argentina to estimate the number of abortions based on a direct counting methodology, focusing specifically on abortions that occurred within the public health subsector of Buenos Aires province. Considering multiple models, the estimations range from 25.50 (model based on direct case counting) to 30.47 (model based on municipalities with reliable records) abortions per 1,000 women of fertile age. Specifically, the model with adjustment for underreporting (Reliable Registering model) yields a value of 29.92 (95% CI: [29.38; 30.47]).

Mario and Pantelines [11] provide different estimates for the number of induced abortions in Argentina. In the year 2000, it was estimated that the annual abortion rate ranged from 40.8 to 49.0 induced abortions per 1,000 women aged 15 to 49 years using the method of hospital discharges. On the other hand, a study by Atucha and Pailles [10] indicated a rate of around 50.0. It is also known that the abortion rate has been reduced in recent years, related in part to the reduction of unintended pregnancy and the number of births in general [28]. In a recent study, Bearak et al. [28] estimate an average of 32 per 1,000 women of fertile age (27 to 37 considering the uncertainty interval 80% and 95%) in Latin America and the Caribbean. In a country-level estimate, the authors indicate a rate of 33 abortions per 1,000 for Argentina. These estimations are consistent with our Realiable Registering model, that falls within the uncertainity interval predicted for Latin America and the Caribbean.

Several factors can alter the estimate and introduce biases. Although abortion is legal in Argentina, the law that regulates it is recent, thus implying a transformation process by the health system, particularly in relation to the existing information systems. Due to the methodology employed and the data sources, underreporting is a major issue. To correct for this, we utilised a set of municipalities where the records were considered to be sufficiently reliable to constitute the model. The problem of under-reporting is common and has been observed in similar studies [12, 13, 27, 28]. Other factors that can alter the results obtained include abortions by residents of Buenos Aires province that occurred outside the province. This is expected to occur primarily with residents of municipalities in Buenos Aires province adjacent to large urban centers, such as Buenos Aires City. It is worth noting that the inverse flow (residents from other districts who had abortions in Buenos Aires province) is detected and not considered within the estimate. Therefore, this is another factor contributing to underreporting in the estimate.

Under-reporting is a pervasive issue in addressing health and social issues, particularly concerning vulnerable and often low-income populations. This problem extends beyond unconventional healthcare practices to include issues such as the search for missing persons [32, 33, 34, 35] and the monitoring of infectious diseases [36]. A robust information system is crucial for capturing and understanding the full scope of these issues. However, it is equally important to delve into the biases that can lead to under-reporting. Recognizing and addressing these biases is essential for developing an effective decision-making system that accurately reflects the needs and realities of these communities.

The study, which is the first to be conducted after the application of the law [2], is also the first that is based in direct counting of cases. This results shows the importance of setting goals in relation to expanding the network of health establishments needed, the quantity of teams, their territorial distribution, as well as for planning the purchase of necessary supplies to guarantee the right throughout the provincial territory. In this regard, future studies on the heterogeneity and impact of access inequalities in the territory of Buenos Aires are pending, which could facilitate the definition of priority areas.

Lastly, note that estimates have a sociocultural impact. They allow for the dimensioning of the event’s magnitude and thereby highlight that abortion is a common event in the lives of people with the capacity to gestate, dispelling the idea that abortion is an infrequent event or one that can be easily eliminated from the lives of women, trans men, and other people with various gender identities who are able to gestate.

## Data Availability

All data produced in the present study are available upon reasonable request to the authors

## Notes

### Competing Interest Statement

The authors have declared no competing interest.

### Funding Statement

This study did not receive any funding

### Author Declarations

Ethics committee of Ministry of Health of the Province of Buenos Aires gave ethical approval for this work

### Summary of Updates

Updating general information, such as a paragraph in the discussion about the importance of studying under-reported cases, and bibliography related to the use of healthcare databases.

## References

[1] Ministerio de Salud de la Provincia de Buenos Aires (2021). Guía de implementación de la interrupción voluntaria del embarazo en la Provincia de Buenos Aires, en el marco de la Ley Nacional Nº 27.610. Disponible en: https://ministeriodelasmujeres.gba.gob.ar/drive/archivos/guiaimplementacionive.pdf

[2] Ley Nacional N° 27.610 de Interrupción Voluntaria del Embarazo (2020). Available on: https://www.boletinoficial.gob.ar/detalleAviso/primera/239807/20210115

[3] Código Penal de la Nación (1921), artículo 86, segundo párrafo, inciso 1 y 2. https://www.argentina.gob.ar/normativa/nacional/ley-11179-16546

[4] Corte Suprema de Justicia de la Nación, “F.A.L. s/ Medida autosatisfactiva”, 13/03/2012, Disponible en: http://www.saij.gob.ar/corte-suprema-justicia-nacion-federal-ciudad-autonoma-buenos-aires-medidaautosatisfactiva-fa12000021-2012-03-13/123456789-120-0002-1ots-eupmocsollaf

[5] Programa Nacional de Salud Sexual y Procreación Responsable (2015). Protocolo para la atención integral de las personas con derecho a la interrupción legal del embarazo. Ministerio de Salud de la Nación. Available on: https://www.argentina.gob.ar/sites/default/files/senaf/materiales-otros-organismos/

[6] Rovere, M. (2016). El Sistema de Salud de la Argentina como campo; Tensiones, estratagemas y opacidades. Revista Debate Público. Reflexión de Trabajo Social, 6(12), 23–41.

[7] Ministerio de Salud de la Provincia de Buenos Aires (2022). Guía de Establacimientos Públicos, Dirección Provincial de Estadística y Salud Digital. Disponible en: https://www.ms.gba.gov.ar/sitios/infoensalud/establecimientos/

[8] Chiara, M., Di Virgilio, M. M., & Moro, J. (2009). “Inequidad (es) en la atención de la salud en el gran Buenos Aires: Una mirada desde la gestión local”. Postdata, vol. 14, n°1, pp. 97–128.

[9] Instituto Nacional de Estadísticas y Censo [INDEC]. (2010) proyecciones de población para la Provincia de Buenos Aires. https://www.indec.gob.ar/.

[10] Aller Atucha, L., & Pailles, J. (1996). La práctica del aborto en Argentina: actualización de los estudios realizados. Estimación de la magnitud del problema. In La práctica del aborto en Argentina: actualización de los estudios realizados. Estimación de la magnitud del problema.

[11] Mario, S., & Pantelides, E. A. (2009). Estimación de la magnitud del aborto inducido en la Argentina. Notas de población.

[12] Singh, S., Shekhar, C., Acharya, R., Moore, A. M., Stillman, M., Pradhan, M. R., … & Browne, A. (2018). The incidence of abortion and unintended pregnancy in India, 2015. The Lancet Global Health, 6(1), e111–e120.

[13] Singh, S., & Wulf, D. (1994). Estimated levels of induced abortion in six Latin American countries. International Family Planning Perspectives, 4–13.

[14] Luzuriaga, J. P., Mársico, F., García, E., González, V., Kreplak, N., Pifano, M., & González, S. (2021). Impact of vaccines against COVID-19 on new SARS-COV2 infections in health care workers of the Province of Buenos Aires. Rev. argent. salud publica, 21–21.

[15] Zamora, R. J. (2021). Analysis of excess, all-cause mortality in a population with health insurance in Argentina, in the context of the covid-19 pandemic. Revista Argentina de Medicina, 9(4).

[16] González, S., Olszevicki, S., Gaiano, A., Salazar, M., Varela Baino, A. N., González Martínez, V., … & Marsico, F. (2023). Efectividad de refuerzos homólogos y heterólogos luego de esquemas primarios con Sputnik V, Astra-Zeneca y Sinopharm durante el período omicron en adultos mayores de 50 años en la provincia de Buenos Aires. Salud publica: revista del Ministerio de salud de la provincia de Buenos Aires, s30087074-kt72juzpm.

[17] González, S., Olszevicki, S., Salazar, M., Calabria, A., Regairaz, L., Marín, L., … Marsico, F. (2021). Effectiveness of the first component of Gam-COVID-Vac (Sputnik V) on reduction of SARS-CoV-2 confirmed infections, hospitalisations and mortality in patients aged 60-79: a retrospective cohort study in Argentina. EClinicalMedicine, 40.

[18] González, S., Olszevicki, S., Gaiano, A., Baino, A. N. V., Regairaz, L., Salazar, M., … & Marsico, F. (2022). Effectiveness of BBIBP-CorV, BNT162b2 and mRNA-1273 vaccines against hospitalisations among children and adolescents during the Omicron outbreak in Argentina: A retrospective cohort study. The Lancet Regional Health–Americas, 13.

[19] Luzuriaga, J. P., Marsico, F., Garcia, E., González, V., Kreplak, N., Pifano, M., & González, S. (2021). Impacto de la aplicación de vacunas contra COVID-19 sobre la incidencia de nuevas infecciones por SARS-COV-2 en PS de la Provincia de Buenos Aires.

[20] González, S., Olszevicki, S., Gaiano, A., Salazar, M., Regairaz, L., Baino, A. N. V., … Marsico, F. (2023). Protection of homologous and heterologous boosters after primary schemes of rAd26-rAd5, ChAdOx1 nCoV-19 and BBIBP-CorV during the omicron outbreak in adults of 50 years and older in Argentina: a test-negative case–control study. The Lancet Regional Health–Americas, 27.

[21] Instituto Nacional de Estadísticas y Censo [INDEC]. (2010) Censo Nacional de Población, Hogares y Viviendas. https://www.indec.gob.ar/.

[22] Instituto Nacional de Estadísticas y Censo [INDEC]. (2010) proyecciones de población para la Provincia de Buenos Aires según sexo y grupos quinquenales de edad. Disponible en : (https://www.indec.gob.ar/indec/web/Nivel4-Tema-2-24-85).

[23] Juarez, F., Singh, S., Garcia, S. G., & Olavarrieta, C. D. (2008). Estimates of induced abortion in Mexico: what’s changed between 1990 and 2006?. International Family Planning Perspectives, 158–168.

[24] Jones, R. K., & Jerman, J. (2017). Abortion incidence and service availability in the United States, 2014. Perspectives on sexual and reproductive health, 49(1), 17–27.

[25] Newcombe, R. G. (1998). Interval estimation for the difference between independent proportions: comparison of eleven methods. Statistics in Medicine, 17(8), 873–890. doi:10.1002/(SICI)1097-0258(19980430)17:8<873::AID-SIM779>3.0.CO;2-I

[26] Holm, S. (1979). A Simple Sequentially Rejective Multiple Test Procedure. Scandinavian Journal of Statistics, 6(2), 65–70. http://www.jstor.org/stable/4615733

[27] Bearak, J., Popinchalk, A., Ganatra, B., Moller, A. B., Tunçalp, Ö., Beavin, C., … & Alkema, L. (2020). Unintended pregnancy and abortion by income, region, and the legal status of abortion: estimates from a comprehensive model for 1990–2019. The Lancet Global Health, 8(9), e1152–e1161.

[28] Bearak, J. M., Popinchalk, A., Beavin, C., Ganatra, B., Moller, A. B., Tunçalp, Ö., & Alkema, L. (2022). Country-specific estimates of unintended pregnancy and abortion incidence: a global comparative analysis of levels in 2015–2019. BMJ Global Health, 7(3), e007151.

[29] Instituto Nacional de Estadísticas y Censo [INDEC]. (2010) Porcentaje de hogares y de población con Necesidades Básicas Insatisfechas (NBI) provincia de Buenos Aires.

[30] Bürkner, P. C. (2017). brms: An R package for Bayesian multilevel models using Stan. Journal of statistical software, 80, 1–28.

[31] Dirección Provincial de Estadística y Salud Digital. (2021) estadísticas vitales y egresos hospitalarios. https://www.ms.gba.gov.ar/sitios/infoensalud/

[32] Marsico, F., Sibilla, G., Escobar, M. S., & Chernomoretz, A. (2024). The Missing Person problem through the lens of information theory. Forensic Science International: Genetics, 70, 103025.

[33] Marsico, F. L., & Caridi, I. (2023). Incorporating non-genetic evidence in large scale missing person searches: A general approach beyond filtering. Forensic Science International: Genetics, 66, 102891.

[34] Marsico, F. L., Vigeland, M. D., Egeland, T., & Piñero, M. H. (2021). Making decisions in missing person identification cases with low statistical power. Forensic science international: genetics, 54, 102519.

[35] Alvarez, E., Obando, D., Crespo, S., Garcia, E., Kreplak, N., & Marsico, F. (2021). Estimating COVID-19 cases and outbreaks on-stream through phone calls. Royal Society open science, 8(3), 202312.

[36] Vigeland, M. D., Marsico, F. L., Pinero, M. H., & Egeland, T. (2020). Prioritising family members for genotyping in missing person cases: a general approach combining the statistical power of exclusion and inclusion. Forensic Science International: Genetics, 49, 102376.

